# Performance of DeepSeek-R1 in Ophthalmology: An Evaluation of Clinical Decision-Making and Cost-Effectiveness

**DOI:** 10.1101/2025.02.10.25322041

**Authors:** David Mikhail, Andrew Farah, Jason Milad, Wissam Nassrallah, Andrew Mihalache, Daniel Milad, Fares Antaki, Michael Balas, Marko M. Popovic, Alessandro Feo, Rajeev H. Muni, Pearse A. Keane, Renaud Duval

**Affiliations:** Temerty Faculty of Medicine, University of Toronto, Toronto, Ontario, Canada; Faculty of Medicine, McGill University, Montreal, Quebec, Canada; Department of Ophthalmology, Centre Hospitalier de l’Université de Montréal (CHUM), Montreal, Quebec, Canada; Department of Software Engineering, University of Waterloo, Waterloo, Ontario, Canada; Department of Ophthalmology, University of Montreal, Montreal, Quebec, Canada; Centre Universitaire d’Ophtalmologie (CUO), Hôpital Maisonneuve-Rosemont, CIUSSS de l’Est-de-l’Île-de-Montréal, Montreal, Quebec, Canada; The CHUM School of Artificial Intelligence in Healthcare (SAIH), Centre Hospitalier de l’Université de Montréal (CHUM), Montreal, Quebec, Canada; Cole Eye Institute, Cleveland Clinic, Cleveland, OH 44195, USA; Department of Ophthalmology and Vision Sciences, University of Toronto, Toronto, Ontario Canada; Retina Division, Stein and Doheny Eye Institutes, Department of Ophthalmology, University of California, Los Angeles, California, United States of America; Department of Biomedical Sciences, Humanitas University, Via Rita Levi Montalcini 4, 20072. Pieve Emanuele-Milan, Italy; Department of Ophthalmology, St. Michael’s Hospital/Unity Health Toronto, Toronto, Ontario, Canada; Institute of Ophthalmology, University College London, London, UK; NIHR Biomedical Research Centre at Moorfields Eye Hospital NHS Foundation Trust, London, UK

**Keywords:** DeepSeek, ChatGPT, artificial intelligence, large language models

## Abstract

**Purpose:** To compare the performance and cost-effectiveness of DeepSeek-R1 with OpenAI o1 in diagnosing and managing ophthalmology clinical cases.

**Study Design:** Cross-sectional evaluation.

**Methods:** A total of 300 clinical cases spanning 10 different ophthalmology subspecialties were collected from StatPearls. Each case presented a multiple-choice question regarding the diagnosis or management of the clinical case. DeepSeek-R1 was accessed through its public chat-based interface, while OpenAI o1 was queried via an Application Program Interface (API) with a standardized temperature setting of 0.3. Both models were prompted using the Plan-and-Solve+ (PS+) prompt engineering method, instructing them to answer multiple choice questions for each case. Performance was calculated as the proportion of correctly answered multiple choice questions. McNemar’s test was employed to compare the two models’ performance on paired data. Inter-model agreement for correct diagnoses was evaluated via Cohen’s kappa. A token-based cost analysis was performed to estimate the comparative expenditures of running each model at scale, accounting for both input prompts and model-generated output.

**Results:** DeepSeek-R1 and OpenAI o1 both achieved an identical overall performance of 82.0% (n=246/300; 95% CI: 77.3-85.9). Subspeciality-specific analysis revealed numerical variation in performance, though none of these comparisons reached statistical significance (p>0.05).

Agreement in performance between the models was moderate overall (κ=0.503, p<0.001), with substantial agreement in Refractive Management/Intervention (κ=0.698, p<0.001) and moderate agreement in Retina/Vitreous (κ=0.561, p<0.001) and Ocular Pathology/Oncology (κ=0.495, p<0.01) cases. Cost analysis indicated an approximately 15-fold reduction in per-query, token-related expenses when using DeepSeek-R1 compared with OpenAI o1 for the same workload.

**Conclusions:** DeepSeek-R1 demonstrates robust diagnostic reasoning and management decision-making capabilities, performing comparably to OpenAI o1 across a range of ophthalmic subspecialty cases, while also offering a substantial reduction in usage costs. These findings highlight the feasibility of utilizing open-weight, reinforcement learning-augmented LLMs as an accessible, cost-effective alternative to proprietary models.

## INTRODUCTION

Artificial intelligence (AI) is increasingly being explored in ophthalmology, particularly for its potential to augment diagnostic precision and support clinical decision-making. Large language models (LLMs), a subset of foundation models trained on extensive textual datasets, boast advanced natural language processing (NLP) capabilities and the ability to engage in complex reasoning tasks.^1–3^ In ophthalmology, LLMs have been tested on their ability to answer board-style multiple-choice questions, analyze clinical cases, and interpret ophthalmic images.^4–9^ Studies evaluating Generative Pretrained Transformer (GPT) 4 and other advanced models have demonstrated a competitive performance with human clinicians in responding to clinical questions in ophthalmology, positioning them as valuable tools for clinical reasoning and knowledge retrieval.^10,11^ However, while these models excel in structured question-answer formats, studies have highlighted limitations in their ability to manage complex, case-based diagnostic reasoning, particularly in multi-step clinical decision-making or ophthalmic image interpretation.^12,13^

DeepSeek-R1, developed by DeepSeek-AI, has gained global attention for its reasoning-centric design, leveraging reinforcement learning (RL) to improve problem-solving capabilities.^14^ Unlike traditional LLMs, which rely heavily on supervised fine-tuning (SFT), DeepSeek-R1 integrates multi-stage training and self-evolution through RL to refine its logical reasoning and decision-making skills. It was designed with a focus on logical inference, real-time problem-solving, and structured reasoning, making it distinct from generative models that primarily excel in content synthesis. A defining characteristic of DeepSeek-R1 is that it is free to use and reuse, democratizing AI development.^15^ DeepSeek-AI reports that it was trained at a fraction of the cost of other leading LLMs, with an estimated $5.6 million compute budget, compared to the $60 million for Meta’s Llama 3.1 405B, $78 million for OpenAI’s GPT-4, and $191 million for Google’s Gemini Ultra.^16,17^

Despite its promising capabilities, DeepSeek-R1’s performance in ophthalmology has not been evaluated. This study represents the first systematic assessment of DeepSeek-R1’s diagnostic accuracy and decision-making effectiveness in ophthalmology, using clinical cases from StatPearls. By benchmarking DeepSeek-R1 against OpenAI’s state-of-the-art model (o1), we aim to evaluate its performance on diagnosis and management of ocular pathologies, and economic feasibility for potential clinical integration. Given the growing interest in AI-driven diagnostic tools, this study provides critical insights into the viability of open-access, reasoning-based LLMs in ophthalmology and their potential role as an alternative path to proprietary AI models for cost-effective, AI-assisted clinical decision-making.

## METHODS

### Data Source

This was a cross-sectional study conducted in accordance with the Strengthening the Reporting of Observational Studies in Epidemiology (STROBE) guidelines.^18^ We also conducted this study in accordance with the transparent reporting of a multivariable model for individual prognosis or diagnosis (TRIPOD)-LLM, which is an extension of the TRIPODLJ+LJartificial intelligence statement that considers the unique challenges of LLM usage in healthcare.^19^ We used 300 cases from an online database of publicly available questions from StatPearls.^20^ These cases covered a broad range of ophthalmology subspecialities and topics. Each case was classified into one of 10 ophthalmology subspecialties, as defined by the American Academy of Ophthalmology’s *Basic and Clinical Science Course*.^21^ These included Cataract/Anterior Segment (n=10), Neuro-ophthalmology (n=31), Oculoplastics (n=33), Pediatric Ophthalmology/Strabismus (n=13), Retina and Vitreous (n=69), Cornea/External Disease (n=27), Glaucoma (n=23), Ocular Pathology/Oncology (n=46), Refractive Management/Intervention (n=34), Uveitis (n=14). There were no cases identified on Clinical Optics and Fundamentals.

### LLM Access and Parameters

We accessed DeepSeek-R1 through its official chat user interface (UI), which provided direct interaction with the model without requiring the Application Program Interface (API).^22^ Unlike API-based implementations, the web platform does not allow for automated batch processing, necessitating manual input for each query. DeepSeek-R1, like other LLMs, uses a temperature parameter to control response variability when given identical prompts. The temperature scale ranges from 0 (producing the most deterministic responses) to 1 (yielding highly creative outputs). However, the platform does not provide direct control over this setting on the website platform, meaning responses were generated using the default configuration.

We accessed OpenAI o1 using the API.^23^ This enabled us to implement customized automated mass prompting techniques via Google Sheets. Additionally, OpenAI o1’s temperature was set to 0.3. While the optimal temperature for our particular application of the model has not been definitively established, our previous research found that a setting of 0.3 resulted in the highest accuracy.^24^

### Prompt Engineering

Our previous analysis identified the Zero-Shot Plan-and-Solve + (PS+) prompt as the most effective approach, leading us to select it for this study. Originally developed by Wang et al., the PS+ prompt instructs the model to break down its task into structured steps, executing them sequentially with detailed guidance.^25^ Each clinical case was presented as a single, text-based prompt without any associated images. The prompt included the complete case description, a multiple-choice question, and available answer choices. The questions followed a standardized format, consisting of one correct answer and three distractor options.

### Statistical Analysis

Performance was calculated as the proportion of correct responses out of the total number of cases assessed. To compare the performance of DeepSeek-R1 and OpenAI o1, McNemar’s test was employed to assess the statistical significance of differences in accuracy between the two models when applied to the same dataset.^26^ This test, designed for paired categorical data, evaluated whether one model consistently outperformed the other. To quantify the level of agreement between the models, Cohen’s kappa (κ) was calculated, providing insight into whether the two models arrived at similar conclusions beyond what would be expected by chance.^27^ Agreement was categorized as the following: 0-0.20 (none to slight agreement), 0.21-0.40 (fair agreement), 0.41-0.60 (moderate agreement), 0.61-0.80 (substantial agreement), and 0.81-1.00 (near perfect agreement). Confidence intervals for accuracy estimates were calculated using the Wilson Score Interval.^28^ All statistical tests were two-tailed, with a 5% significance threshold. Statistical analyses were performed with R version 4.4.2 (R Foundation for Statistical Computing).

## RESULTS

DeepSeek □R1 achieved an overall accuracy of 82.0% n=(246/300 cases; 95% CI: 77.3-85.9), which was identical to OpenAI o1’s accuracy of 82.0% (n=246/300; 95% CI: 77.3-85.9) (p=1.000) (**Table 1**). Both models correctly answered 224 cases (74.7%), incorrectly answered 32 cases (10.7%), and there were an equal number of discordant cases where one model correctly answered a case that the other incorrectly answered (n=22/300, 7.3%) (**Table 2**). Compared to OpenAI o1, DeepSeek×R1 exhibited a numerically higher diagnostic accuracy than OpenAI o1 in 4 of the 10 subspecialties analyzed, including Cornea/External Disease (88.9% vs. 85.2%), Glaucoma (95.7% vs. 87.0%), Retina/Vitreous (82.6% vs. 81.2%), and Uveitis (85.7% vs. 71.4%). However, none of these differences reached statistical significance (p>0.05). Likewise, OpenAI o1 showed higher numerical accuracy values for Cataract/Anterior Segment (70.0% vs. 60.0%), Neuro-ophthalmology (93.5% vs. 87.1%), Ocular Pathology/Oncology (76.9% vs. 69.2%), Oculoplastics (84.8% vs. 81.8%), and Pediatric Ophthalmology (82.6% vs. 80.4%) (all p>0.05). Both models scored the same on Refractive Management/Intervention (73.5%). **Figure 1** compares the performance of both models by subspeciality.

**Table 1.** Performance of DeepSeek-R1 and OpenAI o1 by Subspeciality.

| Model | Subspeciality | Accuracy (%) | 95% CI (%) | p-value |
| --- | --- | --- | --- | --- |
| DeepSeek-R1 | Cataract/Anterior Segment | 60.0 | [31.3, 83.2] | 1.000 |
| OpenAI o1 |  | 70.0 | [39.7, 89.2] |  |
| DeepSeek-R1 | Cornea/External Disease | 88.9 | [71.9, 96.1] | 1.000 |
| OpenAI o1 |  | 85.2 | [67.5, 94.1] |  |
| DeepSeek-R1 | Glaucoma | 95.7 | [79.0, 99.2] | 0.625 |
| OpenAI o1 |  | 87.0 | [67.9, 95.5] |  |
| DeepSeek-R1 | Neuro-ophthalmology | 87.1 | [71.1, 94.9] | 0.625 |
| OpenAI o1 |  | 93.5 | [79.3, 98.2] |  |
| DeepSeek-R1 | Ocular Pathology/Oncology | 69.2 | [42.4, 87.3] | 1.000 |
| OpenAI o1 |  | 76.9 | [49.7, 91.8] |  |
| DeepSeek-R1 | Oculoplastics | 81.8 | [65.6, 91.4] | 1.000 |
| OpenAI o1 |  | 84.8 | [69.1, 93.3] |  |
| DeepSeek-R1 | Pediatric Ophthalmology | 80.4 | [66.8, 89.3] | 1.000 |
| OpenAI o1 |  | 82.6 | [69.3, 90.9] |  |
| DeepSeek-R1 | Refractive Management/Intervention | 73.5 | [56.9, 85.4] | 1.000 |
| OpenAI o1 |  | 73.5 | [56.9, 85.4] |  |
| DeepSeek-R1 | Retina/Vitreous | 82.6 | [72.0, 89.8] | 1.000 |
| OpenAI o1 |  | 81.2 | [70.4, 88.6] |  |
| DeepSeek-R1 | Uveitis | 85.7 | [60.1, 96.0] | 0.500 |
| OpenAI o1 |  | 71.4 | [45.4, 88.3] |  |
| DeepSeek-R1 | Overall | 82.0 | [77.3, 85.9] | 1.000 |
| OpenAI o1 |  | 82.0 | [77.3, 85.9] |  |

**Table 2.** Contingency Table Comparing DeepSeek-R1 and OpenAI o1.

| *p=1.000* |  | DeepSeek-R1 |  |  |
| --- | --- | --- | --- | --- |
|  |  | Correct | Incorrect | Total |
| OpenAI o1 | Correct | 224 (74.7%) | 22 (7.3%) | 246 (82.0%) |
|  | Incorrect | 22 (7.3%) | 32 (10.7%) | 54 (18.0%) |
|  | Total | 246 (82.0%) | 54 (18.0%) | 422 (100.0%) |

**Figure 1.**
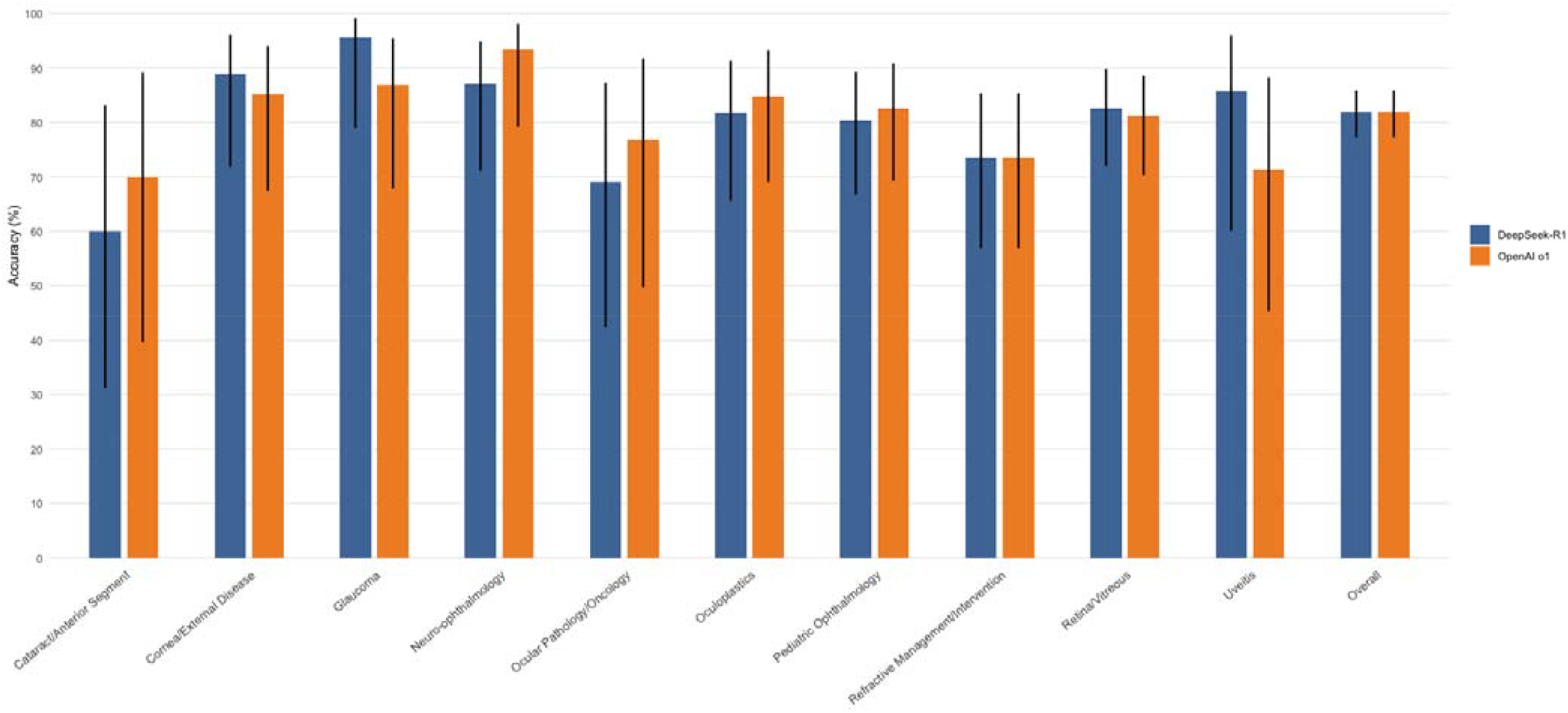
Performance of DeepSeek-R1 and OpenAI by Subspeciality

### Inter-Model Agreement

Cohen’s kappa was calculated to assess agreement between DeepSeek-R1 and OpenAI o1 for correct diagnoses across subspecialties (**Table 3**). Overall agreement across all cases was moderate (κ=0.503, p<0.001). Agreement levels varied by subspecialty, with substantial agreement in Refractive Management/Intervention (κ=0.698, p<0.001) and moderate agreement in Retina/Vitreous (κ=0.561, p<0.001) and Ocular Pathology/Oncology (κ=0.495, p<0.01) cases.

**Table 3.** Agreement between DeepSeek-R1 and OpenAI o1.

| Subspecialty | Cohen's Kappa | p-value |
| --- | --- | --- |
| Cataract/Anterior Segment | 0.783 | 0.067 |
| Cornea/External Disease | 0.509 | 0.069 |
| Glaucoma | -0.070 | 1.000 |
| Neuro-ophthalmology | 0.271 | 0.598 |
| **Ocular Pathology/Oncology** | **0.495** | **0.004** |
| Oculoplastics | 0.238 | 0.457 |
| Pediatric Ophthalmology | 0.418 | 0.411 |
| **Refractive Management/Intervention** | **0.698** | **0.000** |
| **Retina/Vitreous** | **0.561** | **0.000** |
| Uveitis | 0.588 | 0.116 |
| **Overall** | **0.503** | **0.000** |

### Cost-Effectiveness

Across all 300 cases, there were a total of 386,778 characters used to prompt both models. According to OpenAI’s Tokenizer tool, one token is equivalent to approximately 4 characters of text for English text.^29^ Thus, the total number of input tokens used to prompt DeepSeek-R1 and OpenAI o1 was approximately 96,695 tokens. In total, OpenAI o1’s responses were 422,046 characters (105,512 tokens), while DeepSeek-R1’s responses were 855,961 characters (213,990 tokens) when aggregating the responses from all prompts. Under the default pay-per-token rates for OpenAI’s “o1-2024-12-17” model ($15.00 USD per 1×million uncached input tokens and $60.00 USD per 1×million uncached output tokens), these 300 prompts would have cost approximately $7.78 USD in total.^30^ The cost breaks down to roughly $1.45 for input and $6.33 for output. Alternatively, OpenAI offers two monthly subscription packages for its chat UI, which costs $20/month for up to 50 o1 prompts per week or $200/month for unlimited o1 prompts.

By contrast, DeepSeek-R1 is free to use through its public chat UI and mobile app. Thus, we incurred no direct per-query fees during our analysis. DeepSeek also offers an API option for its R1 model, “deepseek-reasoner,” which has a token-based billing model—$0.55 USD per 1×million input tokens and $2.19 USD per 1×million output tokens, including both its chain-of-thought (CoT) and final answer.^31^ Had the API been used, it would have cost $0.52 USD overall. Thus, comparing the estimated API charges, DeepSeek-R1’s cost amounts to 6.71% of OpenAI o1’s total, or a 14.91-fold cost reduction in expenditures at the prompting volume examined in this study.

## DISCUSSION

DeepSeek-R1 demonstrated comparable reasoning capabilities in ophthalmology cases, exactly matching the performance of OpenAI o1 of 82.0% (95% CI: 77.3-85.9) across 300 clinical case multiple choice questions in ophthalmology. Subspecialty-level analysis revealed similar accuracy levels across each subspeciality, with no significant differences in performance between the two models. Despite minor variations, the performance consistency across the subspecialties underscores the capability of both models to handle a broad spectrum of ophthalmic cases. Refractive Management/Intervention, Ocular Pathology/Oncology, and Retina/Vitreous cases revealed moderate to substantial agreement between models (p<0.01). With an overall moderate agreement between the models (κ=0.503, p<0.001), DeepSeek-R1 performed as well as OpenAI o1 while maintaining an approximately 15-fold cost reduction, making it a highly cost-effective yet accurate alternative. Our results align with those of Mondillo et al., who evaluated DeepSeek-R1 in pediatric decision support and noted that while OpenAI o1 achieved slightly higher accuracy (92.8% vs. 87.0%), DeepSeek-R1 demonstrated superior adaptability and accessibility.^32^

In our study, DeepSeek-R1’s comparable performance to OpenAI’s highest performing model can be attributed to its innovative training methodology. Initially, DeepSeek-R1 undergoes a “cold start” phase, where it is trained on a diverse set of carefully selected reasoning datasets before RL to establish a foundational understanding.^14^ DeepSeek-R1 employs a Mixture of Experts (MoE) strategy, which activates specialized reasoning pathways tailored to specific tasks. This approach improves its handling of domain-specific challenges, enabling it to specialize in complex problem-solving across multiple disciplines.^33^ Additionally, DeepSeek-R1 employs rejection sampling and supervised fine-tuning, generating multiple responses to a given problem and selecting only the most accurate and logically coherent outputs for further refinement. This iterative process allows the model to continuously correct mistakes, reinforce high-quality reasoning patterns, and optimize its performance. Prior to its final output, DeepSeek-R1 produces CoT outputs, explicitly discussing its intermediate reasoning steps.

Within these responses, the model autonomously revises its reasoning, recognizing errors and correcting them in real-time. This remarkable self-reflection process, referred to as the “aha moment,” closely resembles human-like cognitive adjustments, allowing the model to iteratively improve its problem-solving skills.^33,34^

Economically, DeepSeek-R1 presents a compelling advantage. Our analysis revealed that, even when utilizing the official API with token-based billing, DeepSeek-R1’s marginal per-query costs are far below those of OpenAI’s o1. It is important to note that DeepSeek-R1 is not fully open-sourced. Instead, it is released under an MIT license as an “open-weight” model, meaning users have access to its pre-trained weights and can build upon its architecture, but the underlying training data remain undisclosed.^17^ Nonetheless, DeepSeek-R1 remains an attractive option for institutions seeking the flexibility of self-hosting a model to reduce costs. While self-hosting may involve infrastructure expenses, the ability to customize and optimize the deployment can lead to long-term savings, particularly for large-scale clinical operations or resource-limited settings. For example, one study evaluated the replacement of OpenAI’s GPT-4 with open-source small language models (SLMs) in a production environment. The research demonstrated that SLMs provided a competitive performance while achieving a cost reduction ranging from 5 to 29 times compared to GPT-4.^35^

One of the key advantages of DeepSeek-R1 is its reasoning-centric design, which could make its decision-making process more interpretable and transparent. Traditional LLMs are often “black boxes,” where users receive an output without insight into how the model arrived at its conclusion. DeepSeek-R1’s CoT feature offers clinicians a transparent view into the model’s reasoning process, allowing them to verify that the logical steps leading to its conclusions are consistent with their own clinical expertise. Nonetheless, this feature does not eliminate the fundamental issue of AI-generated “hallucinations,” defined as instances where the model produces plausible but incorrect medical information. According to Wang et al., these errors pose a significant risk in patient care, particularly when AI outputs influence diagnoses, treatment recommendations, or research conclusions.^36^ Furthermore, the fact that DeepSeek-R1 does not reveal its training corpus means that even when reasoning is made explicit, its responses may still biased by incorrect representations or misrepresentations of medical knowledge.

DeepSeek-R1’s self-reflective capability can be the means by which it bypasses its own safety constraints, which raises security concerns over the model generating outputs that deviate from established medical guidelines or rationalize incorrect treatments that could endanger patient care.^33^ External guardrails such as rule-based reinforcement filters, human-in-the-loop verification, and continuous real-world validation, are essential for safe deployment.^37^ Additionally, retrieval-augmented generation (RAG) techniques and external fact-checking tools could be used to ground the model’s responses in up-to-date, peer-reviewed medical literature.^38^

This study has several limitations that warrant discussion. Firstly, our cost analysis was based solely on the final answers generated by the models, excluding CoT outputs. Since CoT reasoning involves additional tokens to articulate intermediate steps, our approach likely underestimated the actual token usage and associated costs. However, this limitation applied uniformly to both DeepSeek-R1 and OpenAI o1, ensuring a fair comparison. Secondly, we excluded the ophthalmic images from the 21 cases that originally included them, thereby not evaluating multimodal imaging interpretation performance. While both models can handle images, DeepSeek-R1 notes that its image analysis solely extracts text from images. Additionally, we employed the PS+ prompt, which was originally designed to enhance reasoning in models lacking structured problem-solving capabilities, raising the question of whether alternative prompting strategies might yield different results in inherently reasoning-centric models. Furthermore, the publicly available dataset from StatPearls may have been included in the training data of one or both models, leading to potential data leakage. This could artificially inflate model performance by allowing it to recognize or recall previously encountered cases rather than applying genuine clinical reasoning.^39^ However, without direct access to the training corpora, we cannot verify whether the identical high performance of both models resulted from prior exposure to the dataset or from the cases being relatively straightforward and well-aligned with the models’ capabilities. Thus, future research should aim to evaluate these models using larger, more diverse datasets, including those that incorporate ophthalmic imaging, to better assess their applicability in clinical practice. Emerging multimodal AI models, such as Kimi K1.5, have begun integrating vision-language capabilities, offering new possibilities for ophthalmic imaging analysis.^33,40,41^

## CONCLUSION

DeepSeek-R1 demonstrated comparable performance to OpenAI o1 across 300 ophthalmology clinical cases, while also offering a substantial cost advantage. Its enhanced reasoning-centric design appears well suited to a range of clinical scenarios in ophthalmology, supporting its potential as a more accessible AI-driven decision support tool. However, continued research on multimodal prompts and robust external guardrails are necessary to confirm its safety and efficacy in clinical practice.

## Data Availability

All data produced in the present study are available upon reasonable request to the authors.

## ACKNOWLEDGEMENTS

None.

